# Knowledge, Practices, and Determinants of Malaria Prevention in Pregnancy: A Cross-Sectional Study in Weija-Gbawe, Ghana

**DOI:** 10.1101/2025.09.23.25336523

**Authors:** Jeremiah Danso Ampofo, Christopher Ayisah, Divine Tobig Naabil, Emmanuel Boayerik, Elikem Kelly Kyekye, Bismark Peprah Nyantakyi, Loretta Akuamo Dadzie

## Abstract

Malaria in pregnancy remains a significant public health challenge in sub-Saharan Africa, with adverse health outcomes for mothers and foetuses. This study assessed knowledge, attitudes, practices and factors associated with malaria preventive practices among antenatal care attendants in the Weija-Gbawe Municipality, Ghana. A cross-sectional study was conducted among 239 pregnant women attending ANC services. Data were collected using Kobo Collect as a data collection tool and analysed using Stata. Descriptive statistics and inferential statistics including frequencies, percentages, chi-square tests and logistic regression models were employed to identify factors associated with malaria prevention practices. Most participants demonstrated high knowledge, 209 (87.5%) and positive attitudes, 194 (81.2%) toward malaria prevention. However, only 74.5% reported good preventive practices. Only 48 (20%) always slept under the mosquito net. Majority, 165 (69.2%), took anti-malarial drugs to prevent malaria. Over 81 (34%) reported inconsistent use of mosquito nets, and stockouts of antimalarial drugs 29 (12.1%) were cited as barriers. Key factors significantly associated with improved practices included cohabiting marital status (AOR = 4.55; 95% CI: 1.01–20.43), prior malaria testing during pregnancy (AOR = 2.63; 95% CI: 1.09–6.28), and positive attitudes toward prevention (AOR = 7.54; 95% CI: 3.15–18.03). Despite high awareness, gaps persist in translating knowledge into consistent preventive practices. Interventions should prioritise addressing systemic barriers, enhancing partner support, and strengthening education programs to reinforce positive attitudes. These measures are critical to reducing the burden of malaria in pregnancy and achieving maternal health targets in malaria-endemic regions.

## Introduction

Malaria, a potentially fatal disease caused by Plasmodium parasites, remains one of the most significant public health challenges worldwide[1, 2]. Transmitted primarily through the bites of infected female Anopheles mosquitoes, malaria affects millions of people each year, with the most severe consequences observed in vulnerable populations, particularly young children and pregnant women [3]. The latest World Malaria Report indicates a concerning rise in malaria cases globally, from 252 million in 2022 to 263 million in 2023, alongside an estimated 597,000 deaths attributed to the disease [2]. Nearly half of the world’s population is at risk of malaria, highlighting the urgent need for effective prevention and control strategies.

The global burden of malaria is disproportionately concentrated in Africa, where the disease accounts for approximately 94% of all cases and 95% of malaria-related deaths in 2023 [2]. Within this context, children under five years old are particularly at risk, representing nearly 76% of malaria fatalities in the WHO African Region [4]. Notably, eleven African countries contribute to two-thirds of the global malaria burden, underscoring the need for targeted interventions in these high-burden areas. The situation exacerbated in 2020, when malaria-related deaths surged to 627,000, the highest toll recorded in nearly a decade [5]. The two most dangerous species of the malaria parasite, Plasmodium falciparum and Plasmodium vivax, pose significant threats to public health, particularly in sub-Saharan Africa where P. falciparum is predominant [6].

In Ghana, malaria remains a leading cause of morbidity and mortality, accounting for 17.6% of outpatient department visits [7]. Recent studies indicate a high prevalence of malaria among pregnant women, with self-reported rates of at least one malaria episode reaching 76.7% in certain districts of the Ashanti Region [8]. Another investigation found an overall prevalence of 17.1% among pregnant women [9]. The implications of malaria during pregnancy are severe, significantly impacting maternal and neonatal health. Each year, over 125 million pregnant women live in malaria-endemic regions, where malaria is a major preventable cause of adverse pregnancy outcomes [10, 11]. Complications associated with malaria during pregnancy include low birth weight, stillbirth, preterm delivery, congenital malaria, and neonatal anemia [12, 13]. In 2018, approximately 900,000 infants in sub-Saharan Africa were born with low birth weight due to malaria, illustrating the far-reaching effects of the disease [14]. A study conducted in Mali highlighted that malaria infections were responsible for significant percentages of miscarriages, stillbirths, and preterm deliveries among primigravidae [15].

In response to this pressing public health issue, Ghana has made notable strides in malaria control. The country has seen a drastic reduction in malaria-related deaths, from 2,799 in 2012 to just 151 in 2022, while malaria prevalence has decreased from 27.5% in 2011 to 8.6% in 2022 [16]. The Ghana National Malaria Elimination Program has implemented several strategies, including the use of insecticide-treated nets (ITNs) and intermittent preventive treatment with sulfadoxine-pyrimethamine (IPTp), to mitigate the impact of malaria on pregnant women and their infants.

Despite achieving significant coverage of IPTp (80.2%) for the second dose in 2019, the uptake of preventive measures remains inconsistent, with barriers such as delayed antenatal care initiation, inadequate awareness of IPTp dosing schedules, and negative attitudes from healthcare providers hindering progress [17]. Given the critical role of knowledge, attitudes, and practices in shaping health-seeking behaviors, this study aims to assess the understanding of malaria among pregnant women, their adherence to preventive practices, and the factors influencing these behaviors.

## Methods

### Study design

This study adopted a quantitative approach and applied a cross-sectional design. This design was selected due to its effectiveness in assessing prevalence. In this context, the study aimed to examine potential associations between pregnant women’s attitudes and the ANC services they received.

### Study site description

The study was conducted at Weija Gbawe Municipal Hospital, the primary healthcare facility in the Weija-Gbawe Municipality of Ghana’s Greater Accra Region. As per the 2021 population census, the municipality has a population of 213,674, predominantly residing in urban settlements characterised by socioeconomic disparities [18]. The region experiences a tropical climate with seasonal rainfall, fostering mosquito breeding and contributing to year-round malaria transmission. Despite its urban classification, the area faces systemic challenges in healthcare access, including inconsistent availability of insecticide-treated nets (ITNs), antimalarial medications, and limited public health infrastructure.

Weija Gbawe Municipal Hospital serves as the central hub for antenatal care services in the municipality, catering to a large population of pregnant women. Its ANC clinic is a critical entry point for malaria prevention interventions, though operational constraints such as overcrowding, staffing shortages, and resource limitations reflect broader gaps in Ghana’s urban maternal healthcare system. The municipality’s urban environment, marked by dense housing, inadequate waste management, and proximity to stagnant water sources, further elevates malaria exposure risks. These factors collectively position the study site as a representative example of the challenges faced in malaria prevention during pregnancy in resource-constrained urban African settings.

### Study population

The study included pregnant women attending ANC in the Weija Gbawe Municipal Hospital. Pregnant women who visited the Municipal hospital and gave their consent to participate were included in the study. However, pregnant women who were mentally unstable or in unsafe conditions were excluded from the study, as well as those who declined to provide their consent.

### Sample size determination

The sample size for pregnant women in the Weija-Gbawe municipality in the study was determined using the single proportion population formula by Cochran.

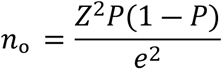

Using Cochran’s formula with an estimated proportion of the population and prevalence of 17% [7], 5% margin of error, and 95% confidence level, a sample of 217 was calculated. To accommodate potential non-responses and incomplete data, an additional 10% was included, resulting in a minimum sample size of 239 participants.

### Sampling procedure

Weija Gbawe Municipal Hospital was deliberately selected through purposive sampling, after which participants were chosen using a simple random sampling method. In this process, eligible individuals were asked to draw from two folded slips labeled “YES” and “NO.” Those who picked “YES” were invited to provide informed consent and participate in the study. If a selected individual declined, another eligible person was randomly chosen, and the procedure was repeated. This continued until the desired sample size was reached, with consideration for possible non-responses during data collection.

### Data collection tool and procedure

Data was collected using the Kobo toolbox with a semi structured questionnaire. The questionnaire comprised four sections covering socio-demographic characteristics of pregnant women, their general knowledge about malaria, and their attitudes and practices toward malaria prevention. The aim was to elicit responses through face-to-face interview techniques.

Before actual data collection, the questionnaire was pretested to ensure its validity and reliability. This step helped identify and make necessary adjustments. Pregnant women were asked to complete the questionnaire, which took approximately 30–45 minutes. For participants who could not read or understand English, trained research assistants administered the questionnaire orally.

Participants were also encouraged to report any ambiguities or difficulties they encountered during the process. Upon completion, the retrieved questionnaires were carefully reviewed, coded, and entered into Stata 17 for data management and analysis. The recruitment period for this study started on 23rd June 2023 and ended on July 18, 2023.

### Ethics approval and consent to participate

The Research Review Committee of the University of Health and Allied Sciences (UHAS-REC) granted ethical approval for the study, with clearance number UHAS-REC A.10[136]22-23. Additional approval was obtained from the Ga-South Municipal Health Directorate and the Weija-Gbawe Municipal Hospital. Prior to data collection, written informed consent was obtained from all participants after they had been fully informed about the purpose of the study. During the consent process, participants were clearly informed that they could leave the study at any time without it affecting the care they received. After the study procedures had been explained, informed consent was obtained from all participants. Those who agreed to participate provided consent by either signing or thumbprinting the consent form. For participants with literacy challenges, a witness was present, and verbal consent was documented when necessary. To ensure confidentiality, only the researchers had access to the responses, which was kept secure under lock and key. Participants were given special codes to prevent their identification by name, and the study’s findings did not include any identifying information. Participants were made aware that their involvement would not provide direct benefits; however, the information collected would be used to generate relevant recommendations.

### Data analysis

The collected data were exported to STATA version 17.0 for cleaning and analysis. Data cleaning involved the removal of duplicates and errors to ensure accuracy. Composite scores for knowledge, attitudes, and practices were generated. For analysis, responses were recorded by grouping correct answers as “yes” and incorrect answers as “no.” A mean score was calculated, and participants scoring below the mean were categorized as having “poor knowledge,” while those scoring above the mean were classified as having “good knowledge.” Descriptive statistics including means or medians, frequencies, and proportions were used to summarize socio-demographic characteristics and KAP variables. Inferential statistics, such as chi-square tests and logistic regression, were employed to assess associations between demographic factors, KAP scores, and malaria prevention practices. Statistical significance was determined at a p-value of less than 0.05, with a 95% confidence interval used to estimate the precision of the results. This analytical approach ensured methodological rigor and helped identify key factors influencing effective malaria prevention among the study population.

## Results

### Demographic Characteristics of Respondents

Table 1 shows that 75 (31.4%) were aged 25-29 years with a mean age of 29.9 (±6.2). More than half 148 (61.9%) were married. Majority 94 (39.3%) of the participants were traders and 41 (17.2%) of the participants were unemployed. Approximately 93 (39%) earned more than GHC 1500 and 30 (12.5%) earned less than 500. Almost two-thirds 158 (66.2%) were Akan’s and 23 (9.6%) were Ga’s. Little above half 128 (53.6%) had given birth before and 111 (46.4%) had not given birth. About 44 (34%) of those who delivered before had two children and 42 (32.8) had one child. All (100%) of them were receiving Postnatal care.

**Table 1.**
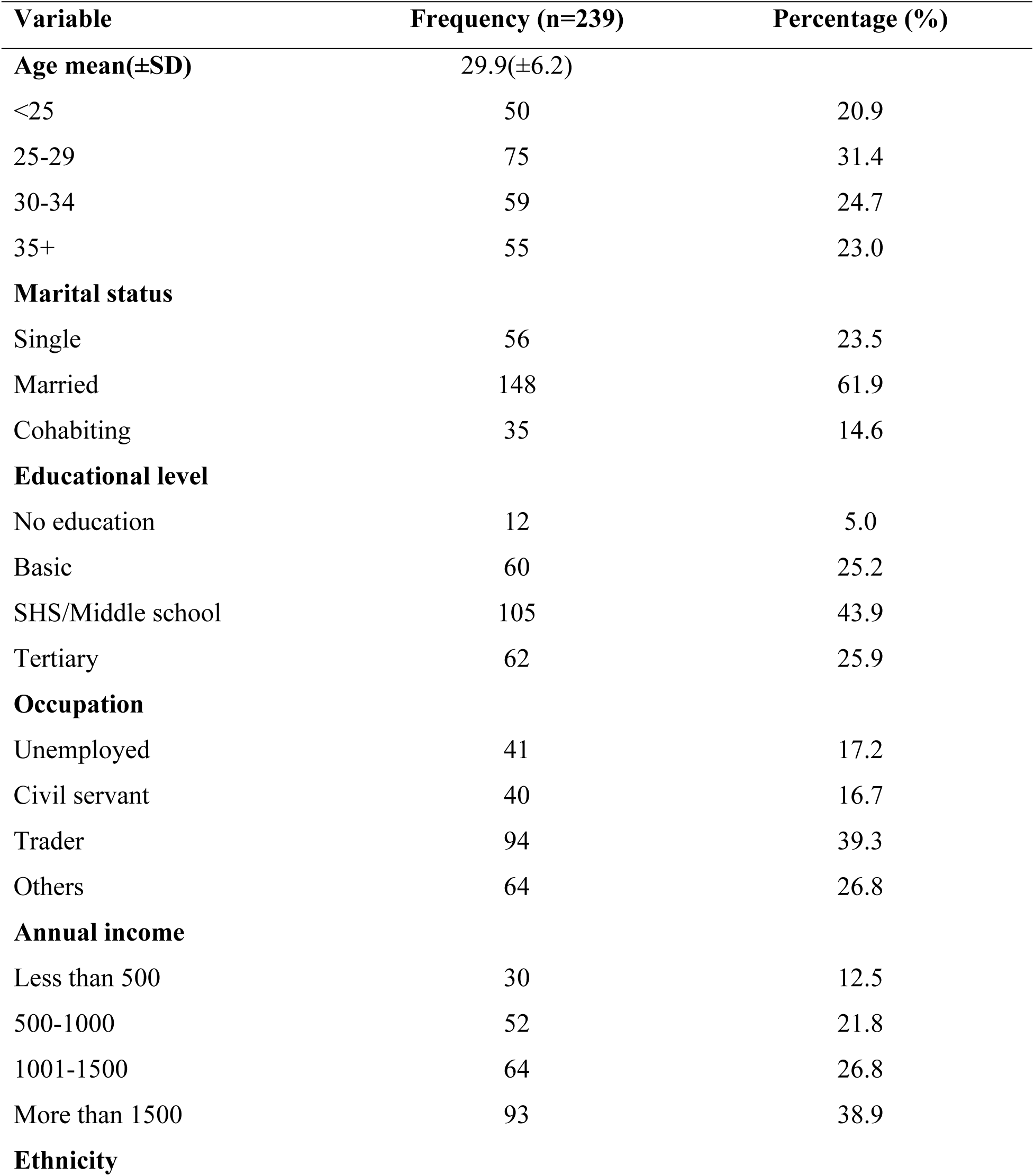
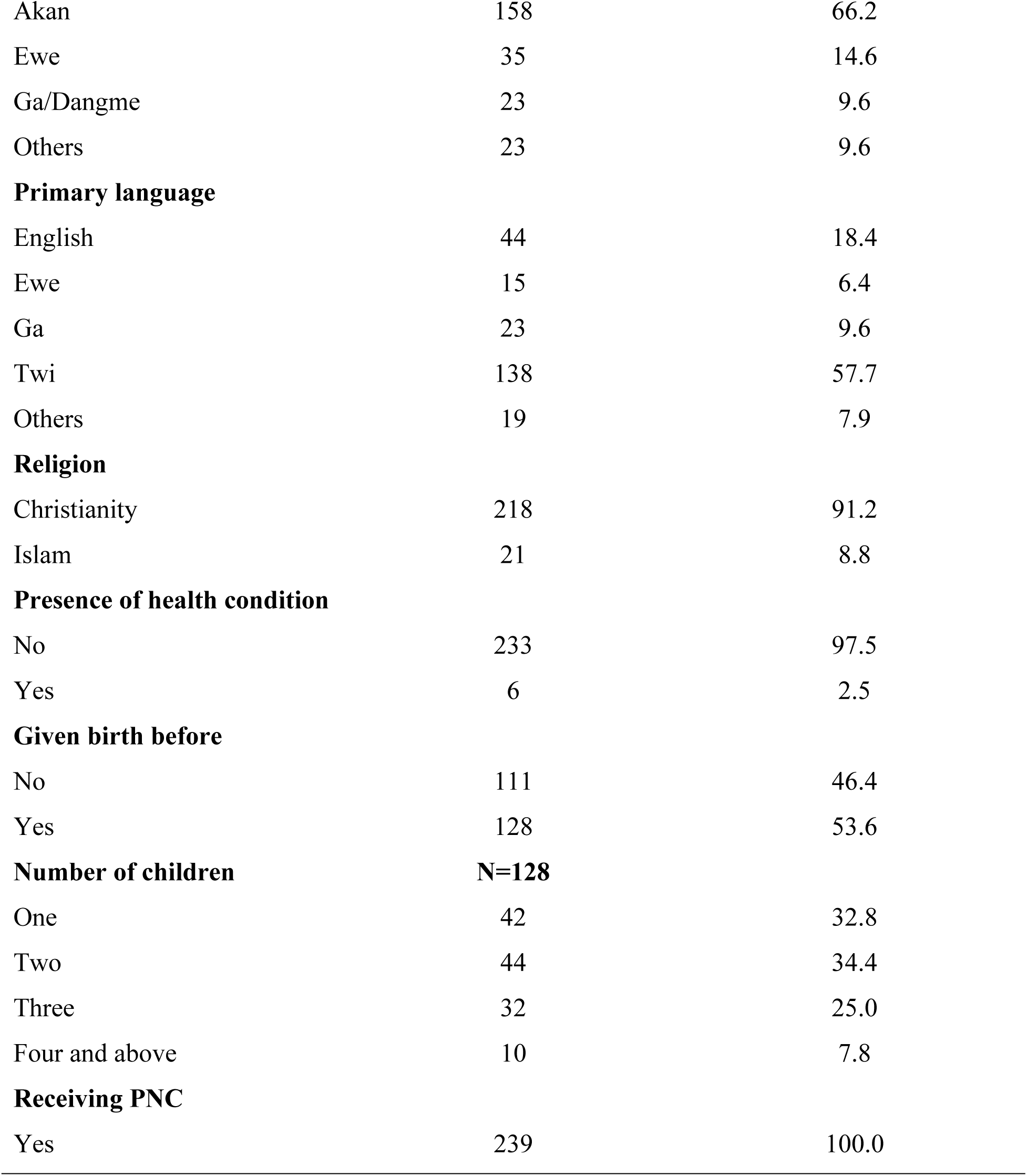
Demographic characteristics of respondents.

### Knowledge About Malaria in Pregnancy

Table 2 presents participant’s knowledge about malaria in pregnancy. Majority, 235 (98.3%) of them had ever heard about malaria. Most, 224 (93.7%) affirmed that pregnant women were at a higher risk of getting malaria. Little above one-third (34.3%) had ever slept under a mosquito during their current pregnancy. All 239 (100%) of them indicated it was important to seek medical attention immediately they experienced symptoms of malaria.

**Table 2.** Knowledge about malaria in pregnancy.

| Item | Frequency<br>(n=239) | Percentage (%) |
| --- | --- | --- |
| Ever heard about Malaria |  |  |
| Yes | 235 | 98.3 |
| No | 4 | 1.7 |
| Pregnant women are at higher risk of getting malaria |  |  |
| Yes | 224 | 93.7 |
| No | 15 | 6.3 |
| Malaria in pregnancy can cause severe complications for both mother and baby |  |  |
| Yes | 230 | 96.2 |
| No | 9 | 3.8 |
| Ever been tested for malaria during your current pregnancy |  |  |
| Yes | 195 | 81.6 |
| No | 44 | 18.4 |
| Taking antimalarial medication can help prevent malaria in pregnancy |  |  |
| Yes | 234 | 97.9 |
| No | 5 | 2.1 |
| Ever taken antimalarial medication during your current pregnancy |  |  |
| Yes | 174 | 72.8 |
| No | 65 | 27.2 |
| Sleeping under a mosquito net can help prevent malaria |  |  |
| Yes | 232 | 97.1 |
| No | 7 | 2.9 |
| Ever slept under a mosquito net during your current pregnancy |  |  |
| Yes | 82 | 34.3 |
| No | 157 | 65.7 |
| Aware it is important to seek medical attention immediately<br>one experiences any symptoms of malaria during pregnancy |  |  |
| Yes | 239 | 100 |
| No | 0 | 0.0 |

### Level of Knowledge Regarding Malaria in Pregnancy

Fig 1 shows that out of the total respondents (239), 209 (87.5%) had a high level of knowledge about malaria in pregnancy, while 30 (12.5%) had low knowledge.

**Fig 1.**
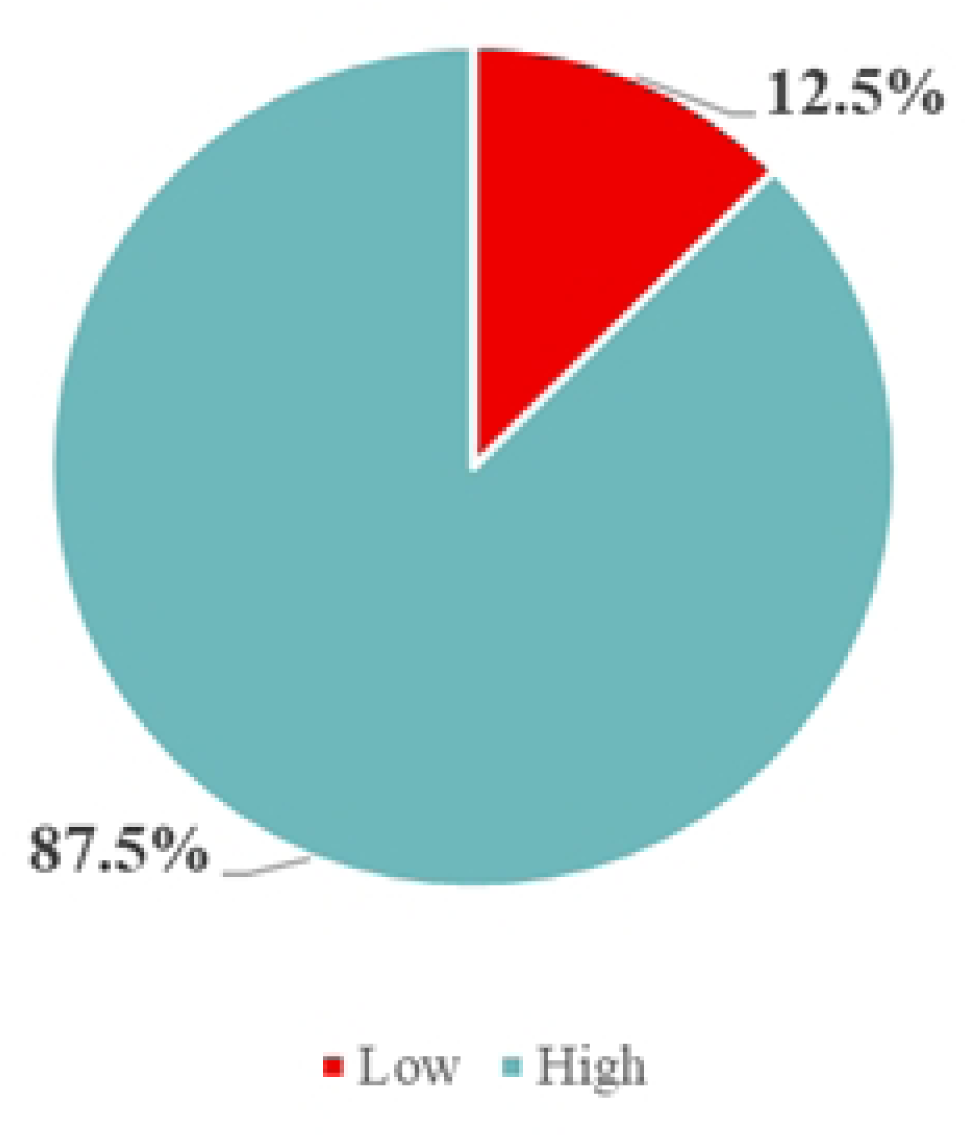
Knowledge Level about malaria in pregnancy.

### Prevalence of Malaria Symptoms

Fig 2 presents the symptoms of malaria as reported by the respondents. The findings show that 232 (97%) experienced chills and fever, 230 (96.2%) reported headache and fatigue, 139 (58.1%) mentioned muscle aches, and 49 (20.7%) experienced diarrhoea and vomiting (Fig. 2)

**Fig 2.**
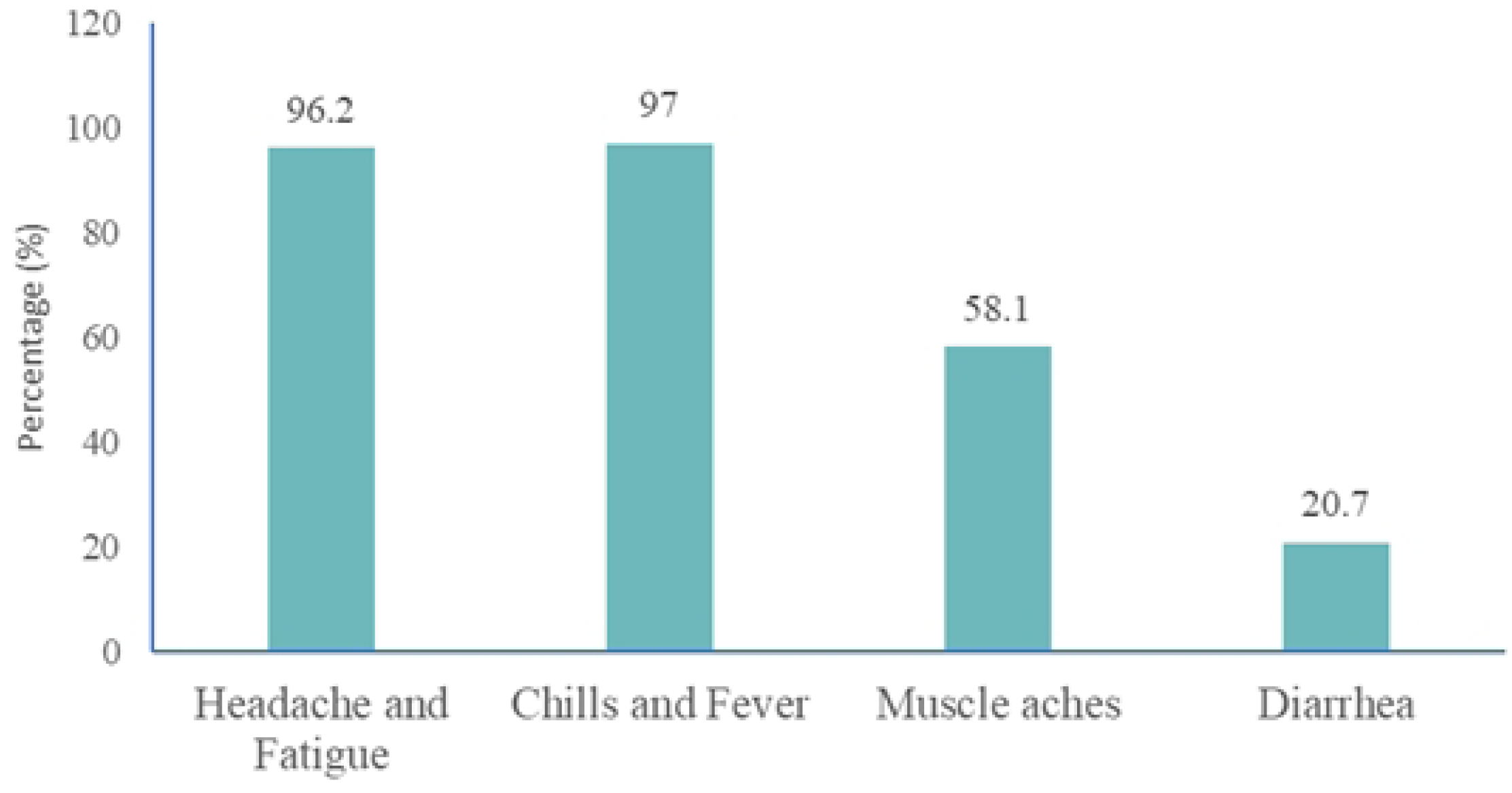
Signs and Symptoms of malaria.

### Attitude Towards Malaria in Pregnancy

Table 3 shows the attitude of study participants towards malaria in pregnancy. More than half, 138 (57.7%) rated the service provider’s attitude as very good. Majority, 237 (99.2%), believed that malaria was a serious health concern during pregnancy. Most 227 (95%) of the participants indicated not experiencing any barrier regarding the usage of malaria prevention tools. More than three-fourths, 213 (89.1%) took preventive steps against malaria during pregnancy. Steps taken were: sleeping under mosquito nets 22 (9.2%) and taking of antimalarial 166 (69.5%).

**Table 3.** Attitude towards malaria in pregnancy.

| **Item** | **Frequency**<br>**(n=239)** | **Percentage (%)** |
| --- | --- | --- |
| **Rate the attitude of service providers towards pregnant women** |  |  |
| Excellent | 17 | 7.2 |
| Fair | 6 | 2.5 |
| Good | 78 | 32.6 |
| Very good | 138 | 57.7 |
| **Services rendered at the Antenatal Care Services** |  |  |
| Health education | 137 | 57.3 |
| Immunization | 1 | 0.4 |
| Screening | 101 | 42.3 |
| **Believe that malaria is a serious health concern during pregnancy** |  |  |
| No | 2 | 0.8 |
| Yes | 237 | 99.2 |
| **Source of motivation to use malaria prevention tools** |  |  |
| Family and friends | 4 | 1.7 |
| Healthcare workers | 8 | 3.3 |
| Importance of prevention methods | 226 | 94.6 |
| Others | 1 | 0.4 |
| **Experiencing barriers hindering usage of malaria prevention tools** |  |  |
| No | 227 | 95.0 |
| Yes | 12 | 5.0 |
| **Important to use malaria prevention tools during pregnancy** |  |  |
| No | 5 | 2.1 |
| Yes | 234 | 97.9 |
| **Taken any preventive measures against malaria during current pregnancy** |  |  |
| No | 26 | 10.9 |
| Yes | 213 | 89.1 |
| **Preventive measures taken by pregnant women** |  |  |
| None | 26 | 10.9 |
| Sleeping under net | 22 | 9.2 |
| Taking antimalarial | 166 | 69.5 |
| Others | 25 | 10.4 |
| **Confidence level based on ability to protect yourself against malaria during pregnancy** |  |  |
| Not confident | 1 | 0.4 |
| Somewhat confident | 23 | 9.6 |
| Very confident | 215 | 90.0 |

### Distribution of Attitude Regarding Malaria in Pregnancy

Fig 3 shows the respondents’ attitudes toward malaria in pregnancy. The results indicate that out of 239 respondents, 194 (81.2%) demonstrated a good attitude, while 45 (18.8%) exhibited a poor attitude.

**Fig 3.**
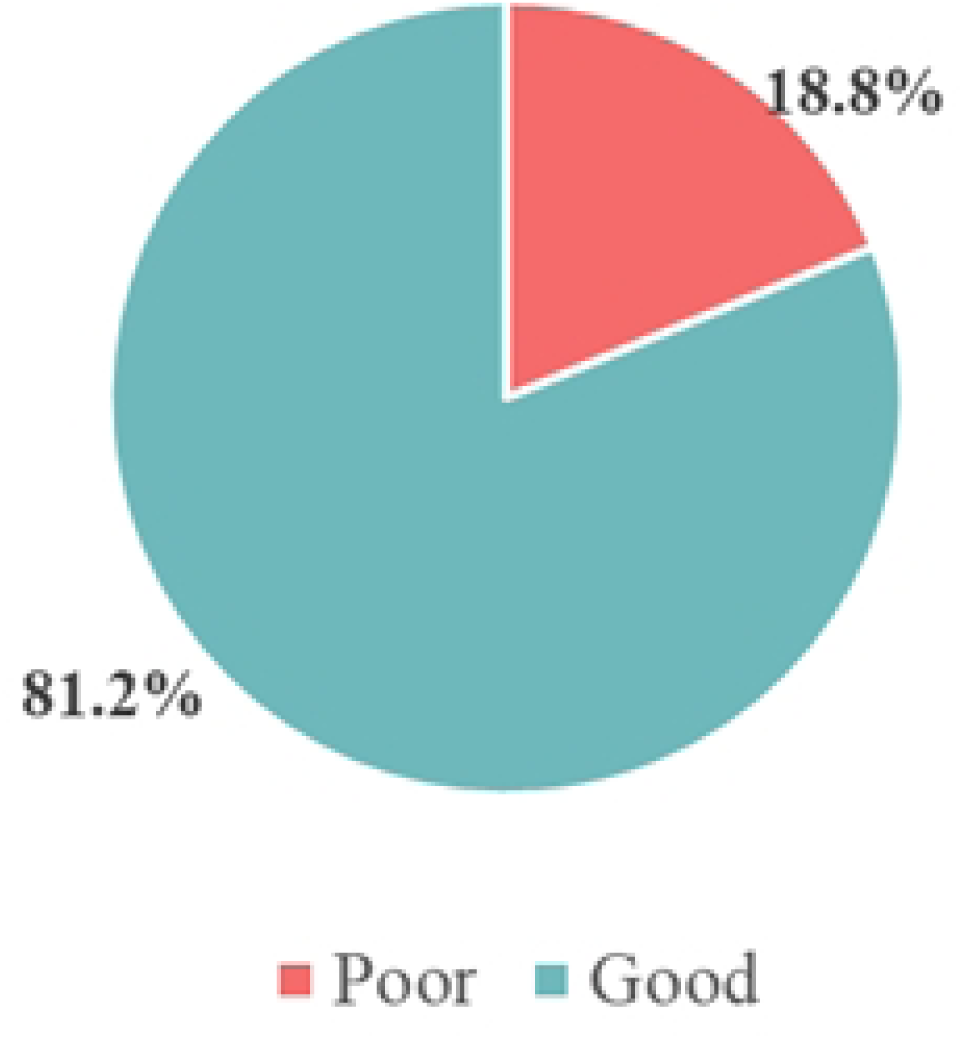
Attitude towards malaria in pregnancy.

### Practices Towards Malaria Preventive Measures in Pregnancy

Table 4 shows practices towards malaria preventive measures in pregnancy. Out of 239 participants, the majority, 186 (77.8%) reported taking malaria preventive medication as prescribed, while 53 (22.2%) did not adhere to their prescribed regimen. Only 11 (4.6%) admitted to ever missing their malaria preventive medication, whereas 228 (95.4%) consistently adhered to the regimen. Regarding malaria incidence, 8 (3.4%) respondents indicated that they had contracted malaria during their current pregnancy, while the overwhelming majority, 231 (96.7%) had not. When asked whether they had received health education about malaria in pregnancy from a health worker, 226 (94.6%) affirmed this, while 13 (5.4%) had not received any such education. In terms of proactive measures, 221 participants (92.5%) had taken steps to prevent malaria during their pregnancy, while 18 (7.5%) had not. Furthermore, 16 women (6.7%) reported experiencing symptoms of malaria during pregnancy, while 223 (93.3%) did not. Among the 16 who experienced symptoms, 15 (93.7%) sought medical attention, whereas only 1 (6.3%) did not.

**Table 4.** Practices towards malaria preventive measures in pregnancy.

| Item | Frequency(n=239) | Percentage (%) |
| --- | --- | --- |
| Take malaria preventive medication as prescribed |  |  |
| Yes | 186 | 77.8 |
| No | 53 | 22.2 |
| Ever missed taking malaria preventive medication |  |  |
| Yes | 11 | 4.6 |
| No | 228 | 95.4 |
| Ever had malaria during this pregnancy |  |  |
| Yes | 8 | 3.4 |
| No | 231 | 96.7 |
| Health education about malaria in pregnancy from a health worker. |  |  |
| Yes | 226 | 94.6 |
| No | 13 | 5.4 |
| Ever taken any steps to prevent malaria during pregnancy. |  |  |
| Yes | 221 | 92.5 |
| No | 18 | 7.5 |
| Ever experienced any symptoms of malaria during pregnancy |  |  |
| Yes | 16 | 6.7 |
| No | 223 | 93.3 |
| Sought for medical attention (n=16) |  |  |
| Yes | 15 | 93.7 |
| No | 1 | 6.3 |

### Malaria Preventive Practices Among Pregnant Women

The majority, representing 178 out of 239 participants (74.5%), exhibited good malaria preventive practices. On the other hand, 61 respondents (25.5%) demonstrated poor preventive practices (see Fig. 4)

**Fig 4.**
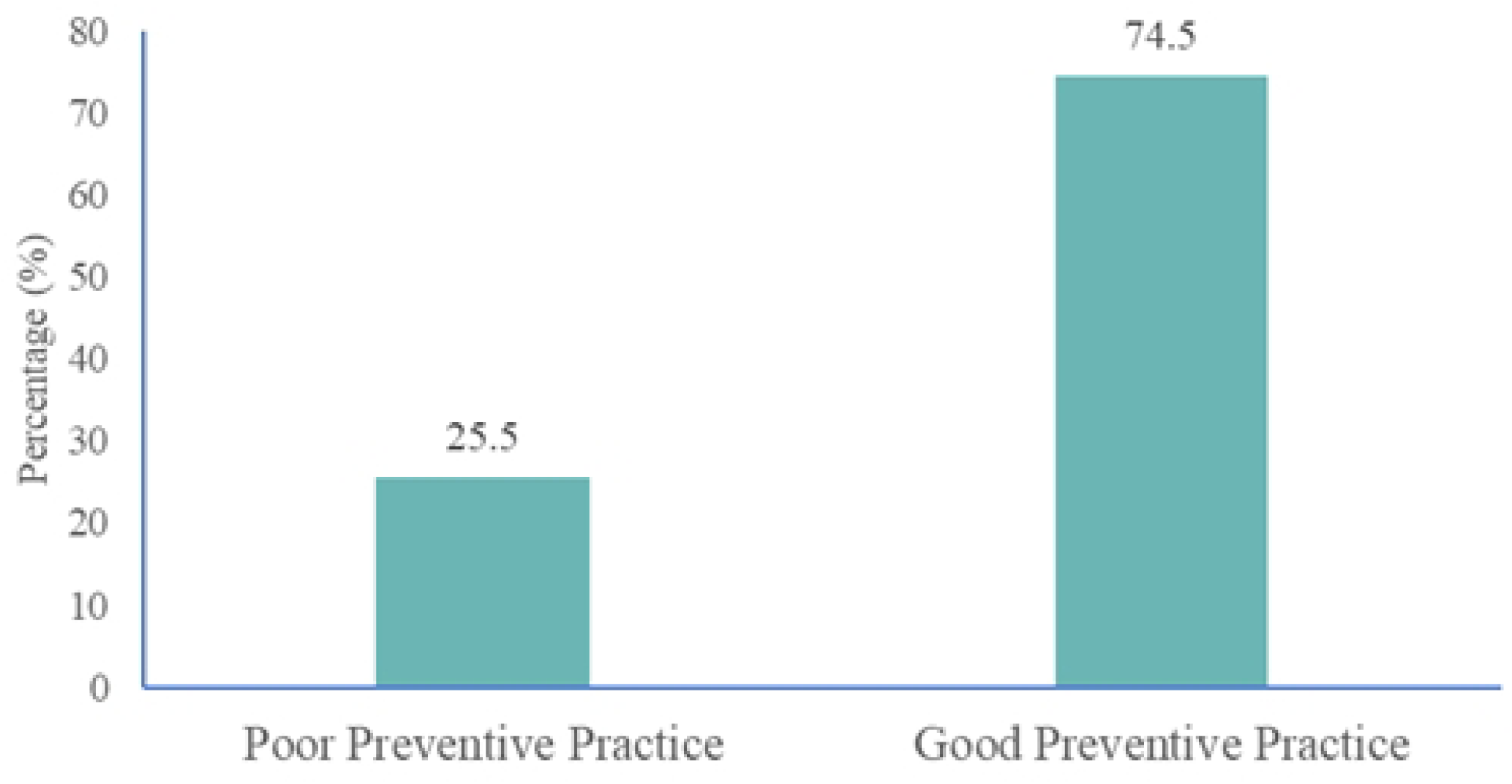
Practices towards malaria preventive measures.

### Frequency of Mosquito Net Use During Pregnancy

Fig 5 illustrates the frequency of mosquito net usage among respondents during their most recent pregnancy. Out of the total participants, the majority, 151 (63.2%), indicated that they never slept under a mosquito net. Meanwhile, 47 (19.7%) reported that they always used a mosquito net, and 41 (17.1%) stated they sometimes used it.

**Fig 5.**
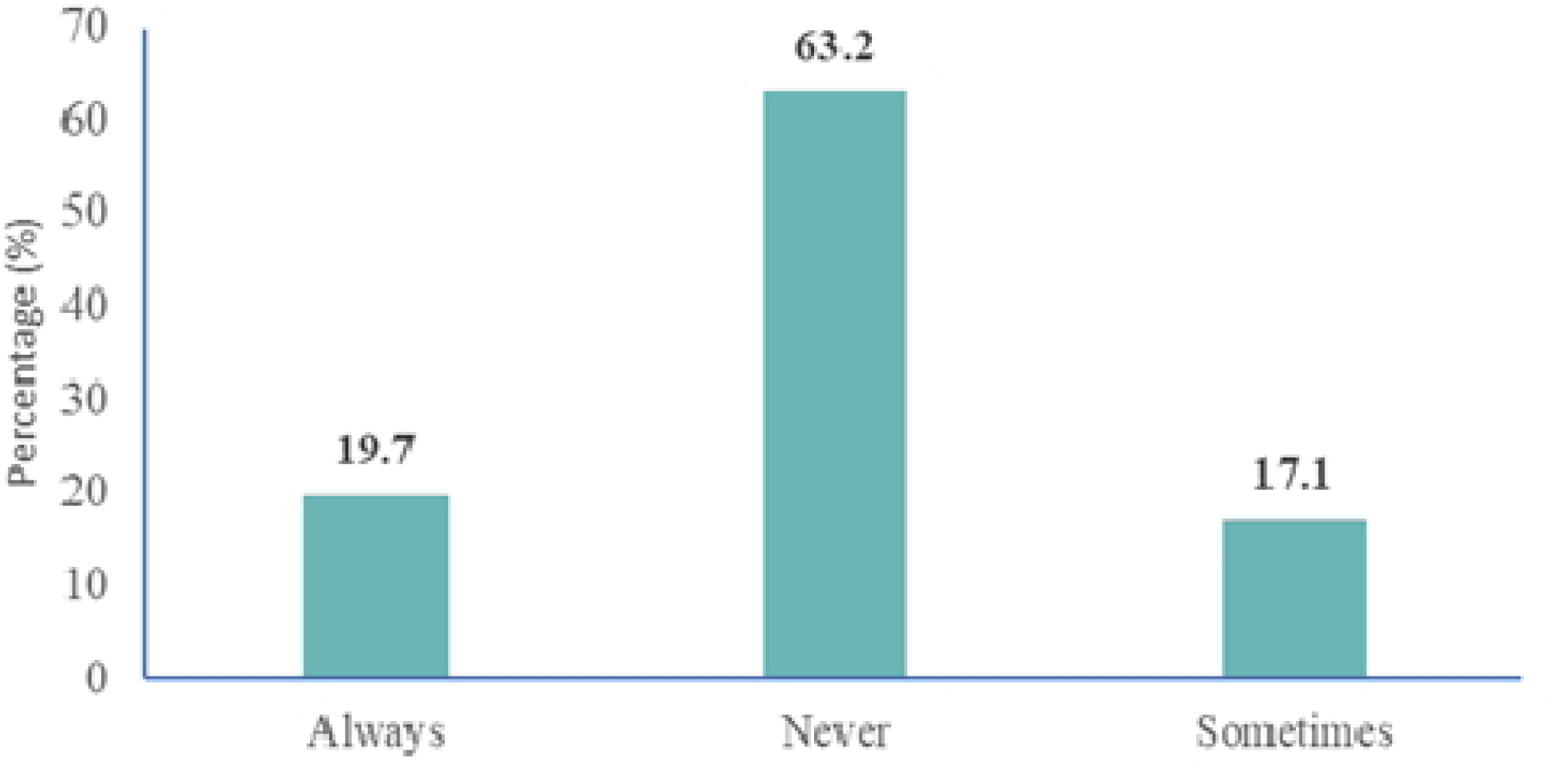
Frequency of sleeping under a mosquito net.

### Utilization of Malaria Preventive Measures During Pregnancy

Fig 6 illustrates the frequency of mosquito net usage among 239 respondents during their recent pregnancy. The majority, 151 (63.2%), reported that they never slept under a mosquito net. About 47 (19.7%) stated that they always used one, while 41 (17.1%) indicated they sometimes did (Fig 6).

**Fig 6.**
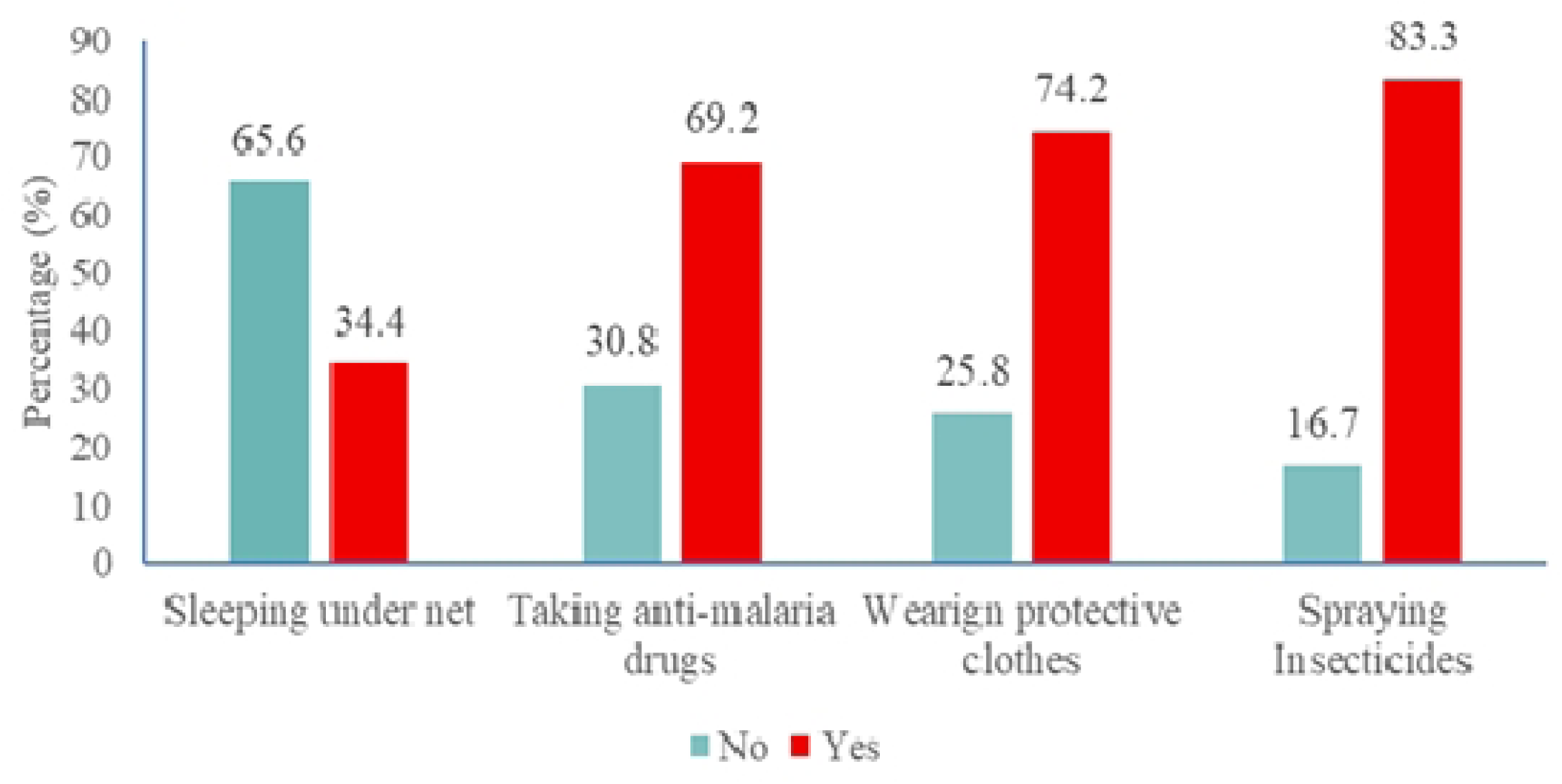
Preventive steps taken to combat malaria.

### Association Between Malaria Preventive Practices and Its Factors

Table 5 shows the Association between malaria preventive practices and its factors. The results revealed that marital status (*χ*2-6.349; p=0.042), educational level of mother (*χ*2=9.254; p=0.026), occupation (*χ*2=9.593; p=0.022), testing for malaria (*χ*2=23.896; p<0.001) and attitude towards malaria (*χ*2=44.181; p<0.001) were significantly associated with the preventive practices.

**Table 5.** Association between malaria preventive practices and its associated factors.

| Variable | Preventive Practices |  | Chi-value (p-value) |
| --- | --- | --- | --- |
|  | Poor 61 (25.5%) | Good 178 (74.5%) |  |
| **Age** |  |  | **0.808(0.848)** |
| <25 | 12(24.0) | 38(76.0) |  |
| 25-29 | 17(22.7) | 58(77.3) |  |
| 30-34 | 17(28.8) | 42(71.2) |  |
| 35+ | 15(27.3) | 40(72.7) |  |
| **Marital status** |  |  | **6.349(0.042)** |
| Single | 17(30.4) | 39(69.6) |  |
| Married | 41(27.7) | 107(72.3) |  |
| Cohabiting | 3(8.6) | 32(91.4) |  |
| **Educational level** |  |  | **9.254(0.026)** |
| No formal education | 6(50.0) | 6(50.0) |  |
| Basic | 19(31.7) | 41(68.3) |  |
| SHS/Middle school | 18(17.1) | 87(82.9) |  |
| Tertiary | 18(29.0) | 44(71.0) |  |
| **Occupation** |  |  | **9.593(0.022)** |
| Unemployed | 12(29.3) | 29(70.7) |  |
| Civil servant | 5(12.5) | 35(87.5) |  |
| Trader | 20(21.3) | 74(78.7) |  |
| Others | 24(37.5) | 40(62.5) |  |
| **Annual income** |  |  | **4.647(0.200)** |
| Less than 500 | 4(13.3) | 26(86.7) |  |
| 500-1000 | 18(34.6) | 34(65.4) |  |
| 1001-1500 | 16(25.0) | 48(75.0) |  |
| More than 1500 | 23(24.7) | 70(75.3) |  |
| **Ethnicity** |  |  | **1.793(0.617)** |
| Akan | 39(24.7) | 119(75.3) |  |
| Ewe | 11(31.4) | 24(68.6) |  |
| Ga/Dangme | 4(17.4) | 19(82.6) |  |
| Others | 7(30.4) | 16(69.6) |  |
| **Religion** |  |  | **0.036(0.850)** |
| Christianity | 56(25.7) | 162(74.3) |  |
| Islam | 5(23.8) | 16(76.2) |  |
| **Presence of health condition** |  |  | **1.939(0.164)** |
| No | 58(24.9) | 175(75.1) |  |
| Yes | 3(50.0) | 3(50.0) |  |
| **Given birth before** |  |  | **0.009(0.922)** |
| No | 28(25.2) | 83(74.8) |  |
| Yes | 33(25.8) | 95(74.2) |  |
| **Ever tested for malaria** |  |  | **23.896(<0.001)** |
| No | 24(54.6) | 20(45.4) |  |
| Yes | 37(19.0) | 158(81.0) |  |
| **Knowledge level** |  |  | 0.024(0.878) |
| Low | 8(26.7) | 22(73.3) |  |
| High | 53(25.4) | 156(74.6) |  |
| **Attitude** |  |  | **44.181(<0.001)** |
| Poor | 29(64.4) | 16(35.6) |  |
| Good | 32(16.5) | 162(83.5) |  |

### Factors Influencing Malaria Preventive Practices Among Pregnant Women

Table 6 shows that participants who were cohabiting were 4.55 times likely to practice good malaria preventive measures compared to those who were singles (AOR=4.55; 95% CI-1.11-20.43). Those who ever tested for malaria during their pregnancy were twice likely to practice good malaria preventive measures compared to those who never tested (AOR=2.63; 95% CI-1.09-6.28). Individuals with good attitude towards malaria prevention techniques were 7 times likely to practice good malaria preventive measures compared to those with poor attitude (AOR=7.54; 95% CI-3.15-18.03).

**Table 6.** Factors influencing malaria preventive practices among pregnant women.

| Covariates | cOR (95% CI), p-value | aOR (95% CI), p-value |
| --- | --- | --- |
| **Age** |  |  |
| <25 | Ref |  |
| 25-29 | 1.08(0.46-2.51), 0.863 |  |
| 30-34 | 0.78(0.33-1.84), 0.571 |  |
| 35+ | 0.84(0.35-2.03), 0.702 |  |
| **Marital status** |  |  |
| Single | Ref | Ref |
| Married | 1.14(0.58-2.23), 0.708 | 1.13(0.48-2.69), 0.775 |
| Cohabiting | **4.65(1.25-17.29), 0.022** | **4.55(1.01-20.43), 0.048** |
| **Educational level** |  |  |
| No formal education | Ref | Ref |
| Basic | 2.16(0.61-7.57), 0.230 | 1.33(0.29-5.93), 0.709 |
| SHS/Middle school | **4.83(1.39-16.71), 0.013** | 3.48(0.80-15.08), 0.095 |
| Tertiary | 2.44(0.69-8.59), 0.164 | 1.74(0.37-8.09), 0.483 |
| **Occupation** |  |  |
| Unemployed | Ref |  |
| Civil servant | 2.89(0.91-9.18), 0.071 |  |
| Trader | 1.53(0.66-3.53), 0.317 |  |
| Others | 0.69(0.29-1.60), 0.87 |  |
| **Annual income** |  |  |
| Less than 500 | Ref | Ref |
| 500-1000 | **0.29(0.09-0.96), 0.043** | 0.30(0.08-1.15), 0.079 |
| 1001-1500 | 0.46(0.14-1.52), 0.205 | 0.34(0.09-1.24), 0.102 |
| More than 1500 | 0.47(0.15-1.48), 0.197 | 0.35(0.9-1.43), 0.145 |
| **Ethnicity** |  |  |
| Akan | Ref |  |
| Ewe | 0.72(0.32-1.59), 0.411 |  |
| Ga/Dangme | 1.56(0.49-4.85), 0.466 |  |
| Others | 0.75(0.29-1.95), 0.555 |  |
| **Religion** |  |  |
| Christianity | Ref |  |
| Islam | 1.11(0.39-3.16), 0.850 |  |
| **Presence of health condition** |  |  |
| No | Ref |  |
| Yes | 0.33(0.07-1.69), 0.184 |  |
| **Presence of barriers in malaria prevention** |  |  |
| No | Ref |  |
| Yes | 1.03(0.27-3.93), 0.966 |  |
| **Given birth before** |  |  |
| No | **Ref** |  |
| Yes | 0.97(0.54-1.74), 0.922 |  |
| **Ever tested for malaria** |  |  |
| No | Ref | **Ref** |
| Yes | **5.12(2.56-10.25), <0.001** | **2.63(1.09-6.28), 0.030** |
| **Knowledge level of malaria in pregnancy** |  |  |
| Low | **Ref** |  |
| High | 1.07(0.45-2.55), 0.878 |  |
| **Attitude towards malaria in pregnancy** |  |  |
| Poor | Ref | **Ref** |
| Good | **9.18(4.47-18.82), <0.001** | **7.54(3.15-18.03), <0.001** |

## Discussion

The findings revealed a high level of knowledge about malaria among the participants, with 87.5% demonstrating good awareness of the disease and its preventive measures. This aligns with previous studies, which reported that 91% of pregnant women were knowledgeable about malaria and its consequences [19]. Consistent with existing literature, a study revealed that majority of pregnant women had good knowledge about the malaria parasite, its transmission, signs and symptoms [20]. Similarly, a study revealed that awareness of IPT-SP was high, with 99.1% of participants reporting they had heard of the intervention [21]. In contrast to this study, a study conducted in East India discovered that a minority of pregnant women, particularly health community workers, were aware of malaria and its treatment [22]. These figures compare favourably with reports from other sub-Saharan African countries, where awareness levels typically range between 70-90% [23]. The high level of knowledge observed in this study may be attributed to the effectiveness of health education programs in ANC clinics, as well as the widespread dissemination of malaria-related information through community health campaigns and efforts of the National Malaria Elimination Program. However, despite this high knowledge, gaps remain in specific areas, such as the consistent use of preventive measures like insecticide-treated nets (ITNs), which were only used by 34.3% of the respondents. This discrepancy between knowledge and practice underscores the need for targeted interventions to bridge this gap.

The study also highlighted positive attitudes toward malaria prevention among pregnant women, with 81.2% exhibiting a good attitude. More than half of the participants rated the attitude of service providers as “very good,” suggesting that positive interactions with healthcare workers may enhance compliance with preventive measures. This affirms findings from other studies, which revealed that 95% of pregnant women exhibited good attitude towards malaria prevention [20]. Similarly, a study revealed that while awareness (99.1%) and compliance (98.8%) were high, only 51.7% achieved optimal IPT-SP uptake [21]. In contrast to this study, a study revealed that majority were aware of the cause of malaria, local name of malaria, mode of transmission, risk of malaria among pregnant women, etc. However, their knowledge and attitude were inadequate regarding the symptomatology and complications of malaria in pregnancy, benefits of sleeping under the net or taking chemoprophylactic doses, or the concurrent use of both [24]. This implies that awareness of malaria may not automatically translate into adequate attitude and practice against malaria but may rather reflect on how individuals perceive various circumstances of using prevention measures and reasons to prevent mosquito bite. The study revealed that most participants received health education about malaria in pregnancy from a health worker. This is similar to a study which revealed that, the dominant role of health providers as the primary source of IPT-SP information underscores their crucial position in health education and service delivery [25]. This finding aligns with previous studies in Nigeria and Ghana that identified healthcare workers as the most trusted source of malaria prevention information [25]. Mothers who got information from health extension workers and TV/radio had significantly better attitude than those who obtained information from neighbours or friends [19]. However, barriers such as financial constraints and limited access to resources were reported by a minority of respondents, indicating that structural challenges persist.

The present study demonstrated a high adherence to malaria prevention among pregnant women, with over 70% following prescribed antimalarial medications and 92.5% engaging in preventive practices. These findings align with a study conducted in Nigeria, which reported increased ITN use and intermittent preventive treatment in pregnancy (IPTp) uptake in both intervention and control groups, with greater improvements observed in the intervention group [26]. However, other studies reported contrasting results. For example, a study conducted in Senegal observed that less than half of the participants owned bed nets, with only about a third actively using them [27]. Most who used the nets reported using them between three to six times per week or nightly, indicating suboptimal and inconsistent usage patterns.

Similarly, a study conducted in rural setting of Ghana found that although mothers attending the Kintampo Municipal Hospital had high knowledge of malaria preventive measures, this knowledge was poorly translated into practice [28]. This discrepancy between knowledge and behavior is supported by findings from a study conducted among urban residents in Yaoundé, which showed that although many individuals were aware of malaria prevention strategies, only a few adhered to them [29]. The variation in results across studies may be attributed to differences in study settings and populations. For instance, the current study was facility-based and focused on pregnant women attending antenatal care, whereas [29] study involved a broader community sample with diverse backgrounds, potentially influencing the observed practices.

Moreover, a study conducted in the city of Yaoundé noted that in many households, ITN use was supplemented with other mosquito control measures such as insecticide sprays, coils, repellents, or window screens, particularly in areas close to mosquito-prone environments like marshlands [29]. Compared to findings from other regions, net utilization among pregnant women in this study was lower than those reported in Tanzania [30], Kenya [31], and Nigeria [32], but higher than in India [33]. Likewise, a study conducted in the Afar Pastoral Region of Ethiopia reported that only 17.3% of participants had good prevention practices, a figure lower than that observed in the present study [34]. This disparity may result from differences in study populations, as our study specifically involved women with a record of antenatal care (ANC) attendance, which often includes malaria prevention education and interventions.

The current study also identified socio-demographic factors influencing preventive practices. Marital status, education level, and attitudes towards malaria prevention were all significantly associated with behavior. Cohabiting women were more likely to adopt preventive measures than single women, possibly due to shared decision-making and support. Furthermore, women who had previously been tested for malaria during pregnancy demonstrated better preventive behavior, suggesting that routine malaria testing in ANC clinics could enhance prevention.

In contrast, a study conducted in the Afar Pastoral Region of Ethiopia found that being married or illiterate was significantly associated with a negative attitude toward malaria prevention [34]. Meanwhile, a study conducted in Senegal highlighted the role of adolescent education level, household head’s education, and household wealth in shaping malaria prevention practices [27]. Adolescents with a positive attitude were nearly twice as likely to engage in preventive behavior than those with a negative outlook. This supports the notion that perception of risk and belief in the effectiveness of preventive strategies are critical motivators of behavior change [35]. When individuals gain sufficient knowledge and develop a favorable attitude toward a disease and its prevention, they are more likely to engage in protective behaviors.

Overall, these findings reinforce the importance of health education, regular ANC attendance, and attitude transformation in improving malaria prevention practices among pregnant women. Tailored community-based interventions that account for individual, socio-economic, and cultural contexts are necessary to bridge the gap between knowledge and practice.

## Conclusion

This study assessed the knowledge, attitudes, and practices of pregnant women regarding malaria prevention in the Weija-Gbawe Municipality. Findings revealed strong awareness, with 87.5% demonstrating good knowledge about malaria risks and preventive measures. Positive attitudes were prevalent, as most respondents recognized malaria as a serious health concern and rated healthcare services favourably. However, gaps existed in practice, while over 70% adhered to antimalarial medication, only 34% consistently used mosquito nets. Key factors influencing preventive practices included marital status, education, malaria testing, and attitudes toward prevention. Social support and resource accessibility also played crucial roles. To improve outcomes, interventions should enhance health education, ensure equitable access to preventive tools, and strengthen collaboration between healthcare providers and communities. Addressing these gaps can reduce malaria-related complications in pregnancy and support broader maternal health goals. Further research should evaluate the long-term impact of such interventions in similar settings.

### Limitation

The cross-sectional design captures data at a single point in time, limiting the ability to observe changes or trends over a longer period. This design also restricts the establishment of cause-and-effect relationships between variables. Moreover, the study did not assess the physical condition or effectiveness of the insecticide-treated nets, such as whether they were treated with insecticide or had any damage. Future research should consider intervention-based designs, such as pre- and post-intervention studies, to assess the impact of health promotion activities on improving knowledge, attitudes, and the consistent use of malaria prevention methods.

## Availability of data and materials

The datasets used in this study are not publicly available due to ethical and confidentiality considerations. However, with approval from the ethics review board and upon reasonable request, the corresponding author can provide anonymized data.

## Acknowledgments

The authors extend their sincere appreciation to the administrative teams of the participating organizations and the study participants for granting permission to undertake the research and for their active involvement, as well as to all individuals who supported the successful completion of this study.

## Authors’ contributions

**Conceptualization**: Jeremiah Danso Ampofo, Christopher Ayisah, Divine Tobig Naabil

**Data curation**: Emmanuel Boayerik, Loretta Akuamo Dadzie, Elikem Kelly Kyekye, Bismark Peprah Nyantakyi

**Formal analysis**: Christopher Ayisah, Divine Tobig Naabil, Emmanuel Boayerik

**Funding acquisition**: Jeremiah Danso Ampofo

**Investigation**: Emmanuel Boayerik, Loretta Akuamo Dadzie, Elikem Kelly Kyekye, Bismark Peprah Nyantakyi

**Methodology**: Jeremiah Danso Ampofo, Christopher Ayisah, Divine Tobig Naabil, Emmanuel Boayerik, Loretta Akuamo Dadzie, Elikem Kelly Kyekye, Bismark Peprah Nyantakyi

**Project administration**: Jeremiah Danso Ampofo, Christopher Ayisah, Divine Tobig Naabil

**Resources**: Jeremiah Danso Ampofo, Christopher Ayisah, Divine Tobig Naabil

**Software**: Christopher Ayisah, Divine Tobig Naabil

**Supervision**: Christopher Ayisah, Divine Tobig Naabil, Emmanuel Boayerik, Loretta Akuamo Dadzie, Elikem Kelly Kyekye, Bismark Peprah Nyantakyi

**Validation**: Christopher Ayisah, Divine Tobig Naabil

**Visualization**: Jeremiah Danso Ampofo, Christopher Ayisah, Divine Tobig Naabil, Emmanuel Boayerik, Loretta Akuamo Dadzie, Elikem Kelly Kyekye, Bismark Peprah Nyantakyi

**Writing – original draft**: Jeremiah Danso Ampofo, Christopher Ayisah, Divine Tobig Naabil, Emmanuel Boayerik, Loretta Akuamo Dadzie, Elikem Kelly Kyekye, Bismark Peprah Nyantakyi

**Writing – review & editing**: Jeremiah Danso Ampofo, Christopher Ayisah, Divine Tobig Naabil, Emmanuel Boayerik, Loretta Akuamo Dadzie, Elikem Kelly Kyekye, Bismark Peprah Nyantakyi

